# Retrospective large-scale evaluation of an AI system as an independent reader for double reading in breast cancer screening

**DOI:** 10.1101/2021.02.26.21252537

**Authors:** Nisha Sharma, Annie Y. Ng, Jonathan J. James, Galvin Khara, Eva Ambrozay, Christopher C. Austin, Gabor Forrai, Georgia Fox, Ben Glocker, Andreas Heindl, Edit Karpati, Tobias M. Rijken, Vignesh Venkataraman, Joseph E. Yearsley, Peter D. Kecskemethy

## Abstract

**Importance:** Screening mammography with two human readers increases cancer detection and lowers recall rates, but workforce shortages make double reading unsustainable in many countries. Artificial intelligence (AI) as an independent reader in double reading may support screening performance while improving cost-effectiveness. The clinical validation of AI requires large-scale, multi-vendor studies on unenriched cohorts.

**Objective:** To evaluate the performance of the Mia^®^ AI system on data that the AI system would process in real-world deployments.

**Design:** A retrospective study simulating the impact of AI on an unenriched screening sample.

**Setting:** Seven European breast screening sites representing four centers: three from the UK and one in Hungary (HU), between 2009 and 2019.

**Participants:** The sample included 275,900 cases (177,882 participants) from seven screening sites, involving two countries and four hardware vendors from 2009 to 2019.

**Intervention:** Simulation of double reading using AI as an independent reader in breast cancer screening on historical data.

**Main Outcomes and Measures:** Performance was determined for standalone AI compared to the historical single reader and for simulated double reading with AI compared to historical double reading, assessing non-inferiority and superiority on relevant screening metrics using a non-inferiority margin of 10% relative difference and a one-sided alpha of 2.5% for both tests.

**Results:** Standalone AI detected 29.8% of missed interval cancers. When compared with historical double reading, double reading with AI showed non-inferiority for sensitivity and superiority for recall rate, specificity and positive predictive value. AI as an independent reader reduced the workload for the second human reader but increased the arbitration rate from 3.3% to 12.3%. Applying the AI system could have reduced the human reading time required by up to 44.8% and reduced the recall rate by a relative 7.7% (from 5.2% to 4.8%).

**Conclusions and Relevance:** Using the AI system as an independent reader maintains or improves the double reading standard of care, while substantially reducing the workload. Thus, it has the potential to provide operational and economic benefits.

**Trial Registration:** Registered on ISRCTN, study ID: ISRCTN18056078

## Introduction

Despite improvements in therapy, breast cancer remains the leading cause of cancer-related mortality among women worldwide, accounting for approximately 600,000 deaths annually (1). Randomised trials and incidence-based mortality studies have demonstrated that population-based screening programs substantially reduce breast cancer mortality (2-6).

Full-field digital mammography (FFDM) is the most widely used imaging modality for breast cancer screening globally (7, 11-14). Using two readers (double reading), with arbitration, increases cancer detection rates by 6-15%, while keeping recall rates low (8-10). The model is standard practice in at least 27 countries in Europe, and in Japan, Australia, the Middle East and the UK (11-14). The high cost of two expert readers to interpret every mammogram, alongside growing shortages of qualified readers, means double reading is difficult to sustain (15-17).

Breast radiology has experience using computer-aided detection (CAD) software to automate screening mammogram analysis, which has been adopted by over 83% of US facilities (18). Recent studies question CAD’s benefit to screening outcomes (19-20). When tested in the United Kingdom National Health Service Breast Screening Programme (UK NHSBSP) as an alternative to double reading, a traditional CAD system reduced specificity with a significant increase in recall rates (21).

Modern artificial intelligence (AI) has emerged as a promising alternative. Recent studies suggest the current generation of AI-based algorithms using deep learning may interpret mammograms at least to the level of human readers (22-26). These included small-scale reader studies (22-24) and larger-scale retrospective studies (24-26) performed on artificially enriched datasets, often involving resampling, to approximate a more representative screening population. The imaging datasets were also significantly skewed towards a single mammography hardware vendor. AI and its potential to positively transform clinical practice on real-world screening populations remains to be confirmed, as also highlighted in a recent systematic review (27).

Rigorous large-scale studies are needed to assess performance of AI in double reading on diverse cohorts of women across multiple screening sites and programmes, and on unenriched screening data representative of populations the AI will process in real-world deployments (28). Such studies should evaluate model performance on images from various hardware vendors, using the most relevant screening metrics. This study aimed to evaluate whether a novel AI system could act as a reliable independent reader while automating a substantial part of the double reading workflow, and to demonstrate standalone performance compared to historical results.

## Methods

### Study design

The AI system was evaluated firstly comparing the AI system’s standalone performance to the historical first human reader, the only guaranteed independent read at all participating sites. Secondly, simulated double reading performance using AI as an independent second reader was compared to historical human double reading.

All comparisons were determined on the same unenriched cohorts. Patient age, screening interval, and method of cancer detection were representative of a real-world screening population. Performance was measured in terms of sensitivity, specificity, recall rate, cancer detection rate (CDR), positive predictive value (PPV), and arbitration rate (rate of disagreement between the first and second readers) (see Supplement, Section 3). A study protocol detailing inclusion/exclusion criteria and target performance metrics was established prior to opening the study.

The statistical analysis plan (see Statistical Methods) was developed and executed by an external Clinical Research Organisation (CRO) (Veristat LLC, supported by Quantics Consulting Ltd). All results presented for the listed metrics are CRO-verified. Other results presented are post hoc.

The study had UK National Health Service (NHS) Health Research Authority (HRA) (REC reference: 19/HRA/0376) and ETT-TUKEB (Medical Research Council, Scientific and Research Ethics Committee, Hungary) approval (Reg no: OGYÉI/46651-4/2020).

### Study population and samples

All analyses were conducted on a consecutive ten-year historical cohort of de-identified cases from seven European sites representing four centers: three from the UK and one in Hungary (HU), between 2009 and 2019. The three UK centers included Leeds Teaching Hospital NHS Trust (LTHT), Nottingham University Hospitals NHS Trust (NUH), and United Lincolnshire Hospitals NHS Trust (ULH). All sites participate in the UK NHSBSP overseen by Public Health England (PHE) and adhere to a three-year screening interval, with women between 50 and 70 years old invited to participate. A small cohort of women between 47 and 49 years, and 71 and 73 years old who were eligible for the UK age extension trial (Age X) were also included (25). The Hungarian center, MaMMa Klinika (MK), involved four sites and corresponding mobile screening units, which follow a two-year screening interval and invite women aged 45 to 65. Across all sites, women outside the regional screening programme age range, who chose to participate as per standard of care (opportunistic screening) were also included. The study population was representative of the screening demographic in the respective countries. Screening cases were acquired from the dominant mammography hardware vendor at each site: Hologic (at LTHT), GE Healthcare (NUH), Siemens Healthineers (ULH), and IMS Giotto (MK).

In total, 304,360 cases were extracted which were compatible 4-view FFDM screening cases. Cases were excluded in three steps, creating unenriched, representative ‘ten-year’ and ‘one-year’ samples (Figure 1A). This resulted in a final cohort of 275,900 eligible cases from 177,882 participants used for analysis, allowing multiple cases per participant. The ‘one-year’ sample (2015) was used for further analysis as a cohort with more complete IC information.

**Figure 1.**
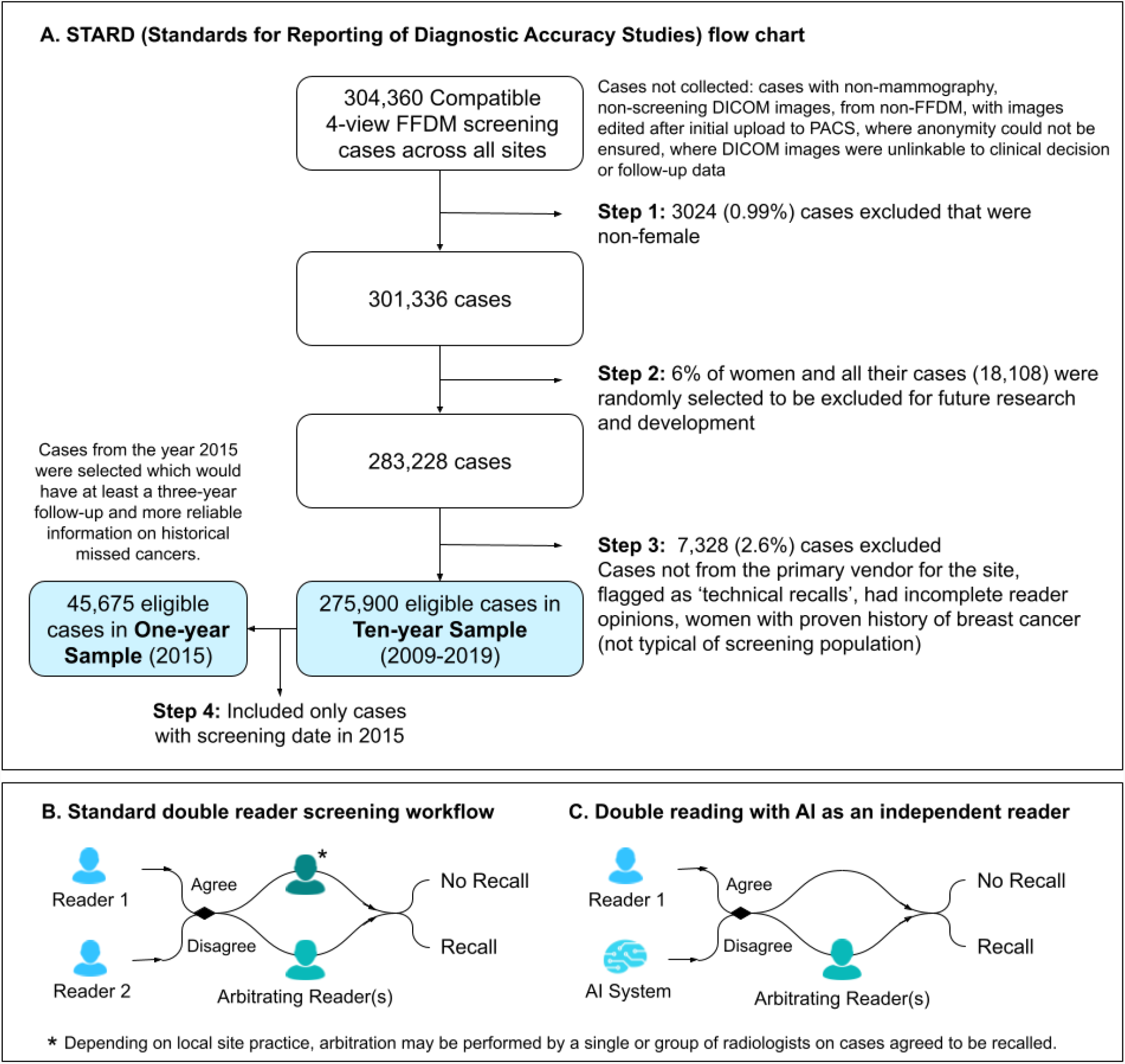
**A**. STARD (Standards for Reporting of Diagnostic Accuracy Studies) flow chart describing case eligibility and the final two study samples, ‘ten-year’ and ‘one-year’. **B**. Standard double reader screening workflow. **C**. Double reading with AI as an independent reader.

### Standard of care double reading and double reading with an AI system

At all sites, the first reader’s opinion was made in isolation, and the second reader had access, at their discretion, to the opinion of the first. In cases of disagreement, an arbitration, performed by a single or group of radiologists, made the definitive “recall” or “no recall” decision. When the opinions agreed “no recall”, a “no recall” decision was reached. When the opinions agreed “recall”, a “recall” decision was reached, or an arbitration performed by a single or group of radiologists made the definitive “recall” or “no recall” decision, depending on the site’s local practice (Figure 1B).

Double reading with the AI system was simulated by combining the opinion of the historical first reader with the AI system (Figure 1C). When both agreed, a definitive “recall” or “no recall” decision was made. Upon disagreement, if available, the historical arbitration opinion was used, otherwise the historical second reader opinion was chosen.

### AI System

All study cases were analysed by the Mia^™^ version 2.0.1 ‘AI system’, developed by Kheiron Medical Technologies. The AI system works with standard DICOM (Digital Imaging and Communications in Medicine) cases as inputs, analyses four images with two standard FFDM views per breast, and generates a binary suggestion of “recall” (for further assessment due to suspected malignancy) or “no recall” (until the next screening interval). The AI system’s output is deterministic, and is based on a single prediction per case. The system used pre-defined thresholds for “recall” or “no recall”.

The AI software version was fixed prior to the study. All study data came from participants whose data was never used in any aspect of algorithm development and was separated from and inaccessible for research and development.

### Determining ground truth, subsample definitions and metrics

All positives were pathology-proven malignancies. All negatives had evidence of a 3-year negative follow-up result. Three-year subsequent cancers include three-year interval cancers (ICs) for the UK plus two-year ICs and additional cancers detected at the next screening round for HU. Recall rate, CDR, and arbitration rate were calculated on the whole population, which included confirmed positives, confirmed negatives, and unconfirmed cases (neither confirmed positive nor negative) as this reflects the real-world screening population. Further details on outcome metrics and ground truthing, including subsample definitions can be found in Supplement, Section 4.

### Statistical methods

A 95% confidence level was used for all confidence intervals (CIs), non-inferiority and superiority testing. Non-inferiority and superiority were tested using relative differences. Non-inferiority was defined to rule out a relative difference of more than 10% in the direction of reduced performance with a one-sided alpha of 2.5%. The 10% margin has been previously used for the assessment of mammography screening with CAD systems, but the 97.5% non-inferiority confidence is stricter than the 90 to 95% commonly used (18). Superiority was tested when non-inferiority was passed and was also based on the same confidence intervals and alpha. Multiplicity was corrected with a gate-keeping method (26-27) for the tests performed on the overall results pooled across regions. See Supplement, Section 6 for further details.

Each vendor (and corresponding study center) had an equal contribution to the observed metrics in this evaluation, for point estimates, confidence intervals and hypothesis tests. Multiple cases were allowed per participant in the ten-year sample, while 99.98% of participants had one case in the one-year sample.

## Results

### Study population and reading workflow

Table 1 presents characteristics of the study population. Of the 275,900 total cases, there were 2792 (1.0%) positives overall (historically detected), made up of 2310 (0.8%) screen-detected positives (in-line with screening expectations) and 482 (0.17%) three-year subsequent cancers (See Supplement, Section 4). For the one-year sample, the percentage of three-year subsequent cancers was significantly higher, comprising 26.0% of all positives (128 out of 493), up from 17.3% (482 out of 2,792) in the ten-year sample. The interval cancer (IC) rates in both the overall and one-year sample were below expectations, which limits the number of positives in the sample (see Supplement, Section 2).

**Table 1:** Characteristics of ten-year and one-year samples.

| Characteristics |  | Ten-year sample<br>(2009-2019) |  | One-year sample<br>(2015) |  |
| --- | --- | --- | --- | --- | --- |
|  |  | Number of<br>cases | Proportion<br>of study<br>population | Number of<br>cases | Proportion<br>of study<br>population |
| **Total<sup>1</sup>** |  | 275,900 | 100.0% | 45,675 | 100.0% |
| Center /<br>Vendor | MK / IMS Giotto | 83,410 | 30.2% | 10,462 | 22.9% |
|  | NUH / GE | 69,045 | 25.0% | 10,983 | 24.0% |
|  | LTHT / Hologic | 64,645 | 23.4% | 10,717 | 23.5% |
|  | ULH / Siemens | 58,800 | 21.3% | 13,513 | 29.6% |
| Age | <40 | 483 | 0.2% | 5 | <0.1% |
|  | 40 - 49 | 37,696 | 13.7% | 5,575 | 12.2% |
|  | 50 - 59 | 114,524 | 41.5% | 19,399 | 42.5% |
|  | 60 - 69 | 98,289 | 35.6% | 16,772 | 36.7% |
|  | 70 - 79 | 23,359 | 8.5% | 3,702 | 8.1% |
|  | 80 - 89 | 1,534 | 0.6% | 221 | 0.5% |
|  | >90 | 15 | <0.1% | 1 | <0.1% |
| Positives | Total positives<sup>2</sup> | 2,792 | 1.01% | 493 | 1.08% |
|  | Screen-detected positives<sup>3</sup> | 2,310 | 0.84% | 365 | 0.80% |
|  | Three-year-subsequent cancer<sup>4</sup> | 482 | 0.17% | 128 | 0.28% |
|  | Three-year ICs from UK<sup>5</sup> | 289 | 0.10% | 80 | 0.18% |
|  | Two-year ICs from HU<sup>5</sup> | 84 | 0.03% | 12 | 0.03% |

See Supplement, Section 1 for annual breakdown of samples and Supplement, Section 4 for more details on ground truthing and subsample definitions.
1. Total number of cases for which CDR, recall rate, and arbitration rate were calculated on (see Supplement, Section 3 and 4 for further details). 2. Used for sensitivity, CDR, and PPV calculations (see Supplement, Section 4 for further details). 3. Screening cases correctly identified in historical double reading, with pathology-proven malignancy (see Supplement, Section 4 for further details). 4. Screening cases not correctly identified in historical double reading, with pathology-proven malignancy arising within 3 years of the screen (see Supplement, Section 4 for further details). 5. Recognising the importance of screening interval differences, these are used for regional analyses (See Supplement, Section 2 and 4 for further details).

### Standalone AI performance

While the AI system is not aimed to operate as a standalone reader in clinical practice, assessing the standalone performance characterises the contribution the AI system could have as an independent reader in the overall double reading workflow. Table 2 presents results for the standalone AI system and the historical first reader. When measuring the AI system performance on historically screen-detected positives without three-year subsequent cancers, the sensitivity was 88.0% (86.7%, 89.3%).

**Table 2:**
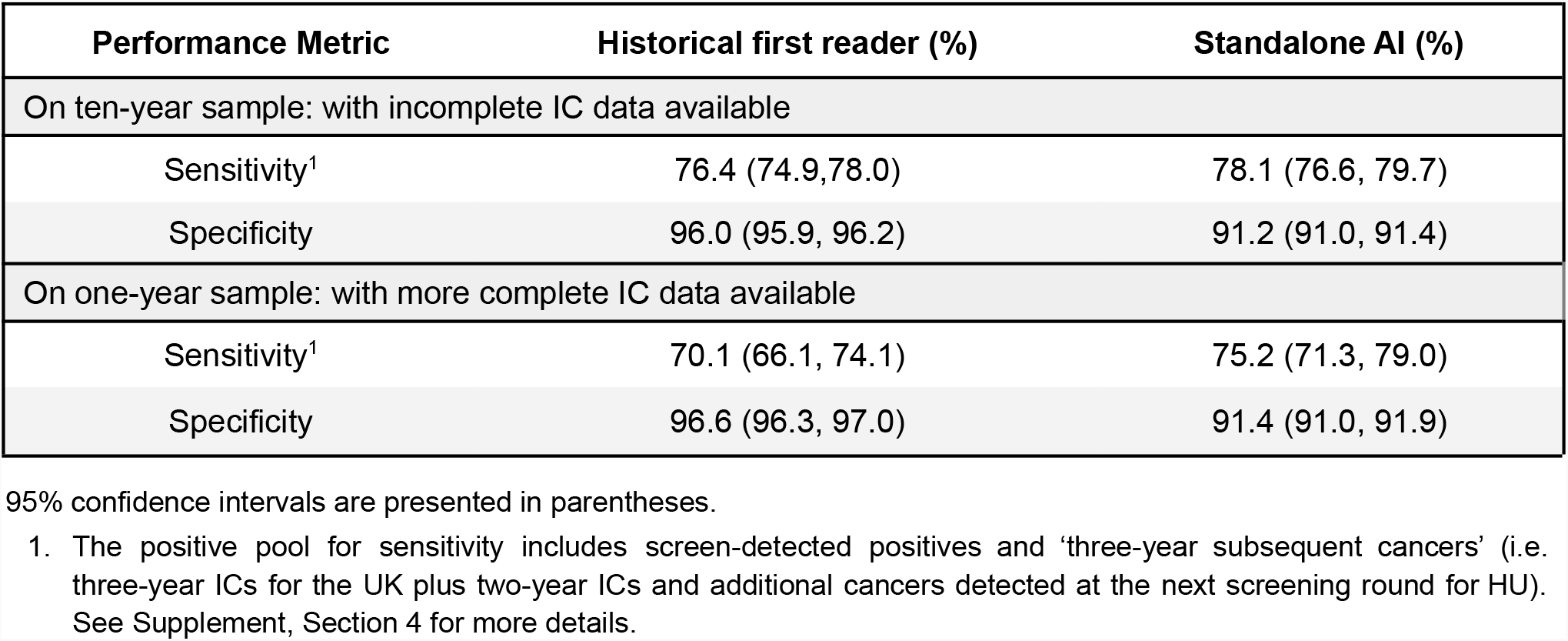
Performance of standalone AI and historical first reader – results pooled across regions.

When compared to historical first reader performance, the AI system showed an absolute difference of 1.7% (0.1%, 3.3%) for sensitivity (including three-year subsequent cancers) and -4.8% (−5.1%, -4.6%) for specificity.

The AI system flagged 2,037 of the 2,310 (88.2%) screen-detected cancers, 111 of the 373 (29.8%) historical ICs (three-year ICs in the UK and two-year ICs in HU), 143 of the 482 (29.7%) historically not detected three-year subsequent cancers, and 177 of 631 (28.1%) cases where cancer was historically detected in the next screening round (3-year screening interval in the UK and 2-year screening interval in HU). In comparison, the historical first reader flagged 2,086 of the 2,310 (90.3%) screen-detected cancers, 26 of the 373 (7.0%) historical ICs, 36 of the 482 (7.5%) three-year subsequent cancers, and 41 of the 631 (6.5%) cases where cancer was historically detected in the next screening round.

Using the one-year sample, where more complete IC data is available, the AI system flagged 46 of the 128 (35.9%) historically not detected three-year subsequent cancers, whereas the historical first reader flagged 6 (4.7%). The AI system also flagged 53 of 198 (26.8%) cases where cancer was historically detected in the next screening round, whereas the historical first reader flagged 9 (4.6%).

### Performance in the double reading workflow

The performance of double reading with AI was estimated using a simulation (see Methods). The statistical tests show that double reading with the AI system compared to historical double reading was at least non-inferior at every metric, with superiority tested and passed for recall rate, specificity and PPV overall (Table 3). Performance by site and vendor are also presented in Supplement, Section 5.

**Table 3:** Performance of double reading with and without AI.

| **A) Results pooled across regions on the ten-year and one-year samples** |  |  |  |
| --- | --- | --- | --- |
| **Performance Metric** | **Historical double reading** | **Double reading (DR) with AI** | **Test outcome for DR with AI<sup>1</sup>** |
| On ten-year sample: with incomplete IC data available |  |  |  |
| Recall rate | 5.2% (5.1, 5.3) | 4.8% (4.7, 4.9) | **Superior** |
| CDR<sup>2</sup> | 8.5 per 1000 (8.4, 8.7) | 8.4 per 1000 (8.2, 8.5) | **Non-inferior** |
| Sensitivity<sup>2</sup> | 84.2% (82.9, 85.6) | 82.4% (81.0, 83.8) | **Non-inferior** |
| Specificity | 96.5% (96.3, 96.6) | 96.8% (96.7, 96.9) | **Superior** |
| PPV<sup>2,4</sup> | 20.5% (20.1, 20.8) | 20.4% (20.1, 20.8) | **Superior** |
| On one-year sample: with more complete IC data available |  |  |  |
| Recall rate | 4.8% (4.6, 5.0) | 4.4% (4.3, 4.6) | **Superior** |
| CDR<sup>2</sup> | 8.1 per 1000 (7.7, 8.5) | 8.0 per 1000 (7.6, 8.4) | **Non-inferior** |
| Sensitivity<sup>2</sup> | 76.1% (72.4, 79.8) | 75.1% (71.3, 78.8) | **Non-inferior** |
| Specificity | 97.0% (96.6, 97.3) | 97.3% (97.0, 97.6) | **Superior** |
| PPV<sup>2,4</sup> | 20.3% (19.3, 21.3) | 20.7% (19.6, 21.8) | **Superior** |
| **B) Regional breakdown on the ten-year sample** |  |  |  |
| **Performance metric** | **Historical double reading** | **Double reading (DR) with AI** | **Test outcome for DR with AI<sup>1</sup>** |
| Regional breakdown for UK |  |  |  |
| Recall rate | 3.8% (3.8, 3.9) | 3.8% (3.7, 3.9) | **Non-inferior** |
| CDR (3Y)<sup>2</sup> | 8.8 per 1000 (8.6, 9.0) | 8.6 per 1000 (8.4, 8.7) | **Non-inferior** |
| Sensitivity (3Y)<sup>2</sup> | 86.1% (84.5, 87.6) | 83.9% (82.3, 85.6) | **Non-inferior** |
| Specificity | 97.1% (96.9, 97.2) | 97.1% (97.0, 97.3) | **Superior** |
| PPV (3Y)<sup>2,4</sup> | 24.5% (24.0, 25.0) | 24.0% (23.5, 24.4) | **Non-inferior** |
| Regional breakdown for HU |  |  |  |
| Recall rate | 9.2% (9.0, 9.4) | 7.8% (7.7, 8.0) | **Superior** |
| CDR (2Y)<sup>3</sup> | 7.7 per 1000 (7.1, 8.3) | 7.6 per 1000 (7.0, 8.2) | **Non-inferior** |
| Sensitivity (2Y)<sup>3</sup> | 88.8% (86.2, 90.9) | 87.5% (84.9, 89.7) | **Non-inferior** |
| Specificity | 94.7% (94.3, 95.0) | 95.8% (95.4, 96.1) | **Superior** |
| PPV (2Y)<sup>3,4</sup> | 8.3% (7.7, 9.0) | 9.6% (8.9, 10.4) | **Superior** |

95% confidence intervals are presented in parentheses.
1. All test outcomes were based on the relative difference with a two-sided 95% CI. A 10% margin was used for non-inferiority testing (see Statistical Methods for details). 2. The positive pool for CDR, sensitivity, and PPV include screen-detected positives and 'three-year subsequent cancers', which are the standard three-year ICs for the UK. 3. The positive pool for CDR, sensitivity, and PPV include screen-detected positives and two-year ICs only, which are relevant for HU. 4. Due to the definition of PPV being over all cases recalled, the figures here represent a lower bound of PPV.

Regional analyses for UK and HU show that at least non-inferiority held for all metrics at both regions well within the 10% margin, with superiority passed for specificity in the UK and superiority passed for RR, specificity, and PPV in HU (Table 3).

### Performance comparison of pathological features

The spectrum of cancers detected by double reading with and without AI was characterised using pre-specified stratification by pathological features for positive cases (Table 4). The maximum absolute percentage difference was 0.7%.

**Table 4:** Pathological features of positive cases recalled in double reading with and without AI.

| Feature | Positive cases from the ten-year sample |  |  |  |
| --- | --- | --- | --- | --- |
|  | Recalled by historical double reading |  | Recalled by double reading first reader + AI |  |
|  | Number of cases | Proportion of positives | Number of cases | Proportion of positives |
| Histological type |  |  |  |  |
| Invasive | 1770 | 75.6% | 1742 | 76.0% |
| In-situ | 345 | 14.7% | 327 | 14.3% |
| Unknown<sup>1</sup> | 226 | 9.7% | 222 | 9.7% |
| Pathological size (invasive only) |  |  |  |  |
| ≤10 mm | 480 | 27.1% | 460 | 26.4% |
| >10 mm | 754 | 42.6% | 750 | 43.1% |
| Unknown<sup>1</sup> | 536 | 30.3% | 532 | 30.5% |
| Lymph node status (invasive only) |  |  |  |  |
| Positive | 364 | 20.6% | 363 | 20.8% |
| Negative | 1263 | 71.4% | 1238 | 71.1% |
| Unknown<sup>1</sup> | 143 | 8.1% | 141 | 8.1% |
| Histology grade (invasive only) |  |  |  |  |
| 1 | 439 | 24.8% | 428 | 24.6% |
| 2 | 937 | 52.9% | 924 | 53.0% |
| 3 | 333 | 18.8% | 330 | 18.9% |
| Unknown<sup>1</sup> | 61 | 3.4% | 60 | 3.4% |

All positives, e.g. screen-detected cancers and 'three-year subsequent cancers' are included.
1. At UK sites, 0.42% of histological type of screen-detected positives were unknown or unavailable.

### Operational performance

When used as an independent reader in a double reading workflow, the AI system automates the second read. This reduction in the number of human readers was offset by an increased proportion of cases requiring arbitration from 3.3% (3.2%, 3.3%) to 12.3% (12.2%, 12.5%) when using the AI system as an independent reader. These results suggest that applying the AI system would have reduced the number of case assessments requiring human readers by 251,914 over the study period. Assuming read times at arbitration may be up to four times greater than first or second reads, this would amount to decreasing the entire workload between 30.0% and 44.8% when accounting for the expected increase in arbitration rate (12.3% vs 3.3%).

## Discussion

To achieve high cancer detection rates while maintaining low recall rates, many European countries rely on double reading, which further exacerbates workforce pressures. An AI system that serves as a robust and reliable independent reader in breast cancer screening addresses both clinical and socioeconomic needs, and helps to make high quality care more widely available. In this large-scale, multi-vendor, retrospective observational study we found that a commercially available AI system could be used as an independent reader in the double reading breast cancer screening workflow.

Double reading performance with the AI, compared to historical double reading, showed superior recall rate (4.8% vs 5.2%) and specificity (96.8% vs 96.5%) and non-inferior cancer detection rate (8.4 vs 8.6 per thousand) and sensitivity (82.4% vs 84.2%). Further, the AI system detected more ICs than the historical first reader (29.8% vs 7.0%), and the comparative cancer detection performance improved when more complete IC data was available. The AI’s sensitivity and CDR performance is limited by the IC data collected (see Supplement, Section 2), therefore, the measured performance in this study is expected to be a lower bound of real-world performance. Importantly, the spectrum of cancers detected in double reading with AI did not change from historical screening results (Table 4), indicating that the use of AI does not require downstream changes to the existing clinical pathway. The reduction in the workload between 30% to 44.8% would significantly lighten the demand for the limited qualified workforce, and may reduce the pressure on screening services.

When assessed on its own, the AI system showed an absolute 1.7% to 5.1% improvement on sensitivity and found 30% to 36% of historical ICs, indicating that cancer detection could be significantly improved with the AI system. The specificity of the AI system was lower than the historical first human reader, which contributed to increased arbitration in double reading. This could potentially be addressed with future improvements of the AI by taking into account the image information available in prior screening rounds which may increase the AI’s specificity.

Past studies have compared the performance of AI systems to individual human readers (22-26). Some employed small-scale reader studies (22-24) with enriched samples of 320 to 720 cases, and larger retrospective evaluations (25-26) with 8,805 to 28,853 cases. While reported performances in small reader studies are encouraging, it remains to be seen if results on enriched test sets and samples generalise to real-world screening populations. Only Kim et al (23) evaluated performance on multiple vendors. McKinney et al (25) demonstrated non-inferiority on both sensitivity and specificity when simulating double reading with an AI system, while Salim et al (26) showed an AI system paired with a single human reader (without arbitration) detects more cancers than two human readers at the cost of significantly higher recall rates. 95% to 100% of cases in both evaluations came from a single hardware vendor, and Salim et al (26) required resampling to approximate a screening population.

The strength of this study is that the AI system was evaluated in simulated double reading on a diverse, heterogeneous, large-scale and representative screening population with data collected across two national screening programmes with a variety of demographic differences. The authors believe this is the first large-scale retrospective study that does not rely on data-construction to approximate a screening cohort, where confirmed positives and negatives, and unconfirmed cases were all included. In contrast, previous studies (32-34) assessed AI performance utilizing cohorts that were enriched for cancer cases and excluded unconfirmed cases, typically the hardest for AI to assess correctly. This is significant as data-construction can introduce unwanted biases and is not guaranteed to faithfully represent a target population or accurately assess AI performance in a real-world environment. The historical reader results represented the practical standard of care, with no influence on reader behavior resulting from participation in the study and no enrichment for positives or any subgroups.

The retrospective nature of the evaluation means a number of obvious limitations. In the simulation, the historical second reader opinion was used as the arbitrator when the historical arbitration opinion was unavailable. This is a lower-bound approximation as arbitrators are informed with previous reader opinions and therefore are expected and have shown to have higher performance than the historical second reader on arbitrated cases. Furthermore, the observed proportion of ICs to positives in the ten-year and one-year samples was lower than expected, although the one-year sample was more complete. With more complete IC data, the point estimate for sensitivity and CDR on the one-year sample can be expected to be more representative, but the smaller cohort size resulted in wider confidence intervals. Estimating the impact of incomplete IC data on outcome metrics will be the subject of future work.

The impact of the historical second reader having discretionary access to the first reader opinion as part of their normal practice may also need further investigation.

While this study demonstrated efficacy in sites already employing double reading, the results suggest the performance standards of double reading could be achieved in programmes currently employing single reading, with lower resources required.

The results from the retrospective evaluation suggest that the AI system could be a promising solution when acting as an independent reader in the double reading workflow. In the simulation, the standard of care was preserved on all relevant screening metrics for double reading comparisons. The scale and diversity of samples provides confidence that the results may generalize to other screening programmes and the use of clinically relevant metrics aims to reliably estimate the impact of introducing AI into everyday screening.

Reducing the overall double reading workload can enable staff redeployment and service improvements such as increased patient interaction, more time for training, an extended programme age range, more focus on complex cases and, during a time of workforce crisis, supporting the sustainability of breast cancer screening.

## Data Availability

The data for the current study are not publicly available. Due to reasonable privacy and ethical concerns, the imaging data cannot be distributed to researchers without ethical approval and research agreements with the original data providers.

## Acknowledgements

We thank M. Bidlek, K. Borbély, G. Di Leo, R. Fülöp, K. Giese, F. Gilbert, T. Helbich, S. Hofvind, B. Joe, K. Keresztes, E. Kovács, M. Milics, Z. Pentek, É. Szabó, L. Tabar, T. Tasnádi, C. Yau for expert input and guidance.

We thank D. Dinneen, S. Kerruish, G. Mehmert, Rianna Mortimer, Cary Oberije, D. Pribil and F. van Beers for technical and management support.

We thank the staff at MaMMa Egészségügyi Zrt., the EMRAD Imaging Network, Nottingham University Hospital Trust and Breast Screening Programme (BSP), The Leeds Teaching Hospital and Leeds/Wakefield BSP, and United Lincolnshire Hospital Trusts and BSP.

Annie Y. Ng and Peter K. Kecskemethy had full access to all the data in the study and take responsibility for the integrity of the data and the accuracy of the data analysis.

Vignesh Venkataraman, Galvin Khara, Tobias M. Rijken, Joseph E. Yearsley, and Georgia Fox conducted and were responsible for data analysis.

## Notes

### Competing Interest Statement

All authors have completed the ICMJE uniform disclosure form. Funding for the UK arm of the study was received from Innovate UK via an NHS England and Improvement, Office of Life Sciences (OLS) Wave 2 Test Bed Programme and a Medical Research Council (MRC) Biomedical Catalyst award. Authors affiliated with Kheiron Medical Technologies are paid employees. There are no financial relationships with any organizations that might have an interest in the submitted work in the previous three years and there are no other relationships or activities that could appear to have influenced the submitted work.

### Funding Statement

The study was funded by Kheiron Medical Technologies. The following authors are employees of the company: Annie Y Ng, Galvin Khara, Georgia Fox, Ben Glocker, Edit Karpati, Tobias M Rijken, Joseph E Yearsley, Peter D Kecskemethy. The following were employees of Kheiron at the time of the study: Christopher C Austin, Andreas Heindl, Vignesh Venkataraman. Eva Ambrozay and Gabor Forrai are paid consultants of Kheiron. All other authors received no payment for this work.
The UK arm of the study was supported by funding from Innovate UK via an NHS England and Improvement, Office of Life Sciences (OLS) Wave 2 Test Bed Programme and a Medical Research Council (MRC) Biomedical Catalyst award.

### Author Declarations

The study had UK National Health Service Health Research Authority (REC reference 19/HRA/0376) and ETT-TUKEB Medical Research Council, Scientific and Research Ethics Committee, Hungary approval (reg no OGYEI/46651-4/2020).

### Summary of Updates

Format of abstract revised. Figure 1 revised. Additional results included in "Standalone AI Performance" section. Discussion revised. Minor typos corrected. Georgia Fox included as author. Supplemental files updated.

