## Supplemental material for "Retrospective large-scale evaluation of an AI system as an independent reader for double reading in breast cancer screening"

### Supplementary Information

#### Table of Contents

**Section 1: Annual breakdown of case numbers** **2**

**Section 2: Regional interval cancer (IC) breakdown** **3**
Table S2: Regional breakdown of historical IC data available compared with expectations 3

**Section 3: Performance metrics** **4**
Table S3: An assessment of metrics used for evaluating performance with AI in breast screening 4

**Section 4: Determining ground truth, subsample definitions and metrics** **6**

**Section 5: Standalone and double reading performance by site and vendor** **7**
Table S4: Performance of standalone AI the historical first reader – by site and vendor. 7

**Section 6: Further statistical details** **10**

**Supplementary Information References** **11**

#### Section 1: Annual breakdown of case numbers

**Table S1: Annual breakdown of case numbers in the entire (ten-year) sample**

| Characteristics | | Ten-year sample (2009-2019) | |
|---|---|---|---|
| | | Number of cases | Proportion of study population |
| **Total** | | 275,900 | 100.0% |
| Year | 2009 | 705 | 0.3% |
| | 2010 | 3,063 | 1.1% |
| | 2011 | 3,179 | 1.2% |
| | 2012 | 13,826 | 5.0% |
| | 2013 | 23,377 | 8.5% |
| | 2014 | 38,523 | 14.0% |
| | 2015 | 45,675 | 16.6% |
| | 2016 | 47,982 | 17.4% |
| | 2017 | 52,160 | 18.9% |
| | 2018 | 44,465 | 16.1% |
| | 2019 | 2,945 | 1.1% |

#### Section 2: Regional interval cancer (IC) breakdown

Table S2 indicates that based on the interval cancer rate (ICR) and proportion of IC numbers observed in the data, IC information is likely incomplete for both the ten-year and the one-year samples, with the one-year sample being more complete.

**Table S2: Regional breakdown of historical IC data available compared with expectations**

| Region | Interval cancer rate (per 1000) | | | Proportion of ICs out of positives (%) | | |
|---|---|---|---|---|---|---|
| | Ten-year Sample | One-year Sample | Expected | Ten-year Sample | One-year Sampled | Expected |
| UK | 1.5 | 2.27 | 3-3.7[1,2] | 14.7 | 21.7 | N/A |
| HU | 1.01 | 1.15 | N/A | 11.7 | 13.5 | 17-20[2] |

The ICs in the UK are defined with their standard three-year interval, and for HU with their two-year interval.

1. The expectation of ICR is based on UK national benchmarks and past studies (1,2).
2. The expectation for the proportion of ICs of positives is based on the requirements of the National Protocol for Breast Cancer Screening in HU (3). The HU center's expectation is 17%.

#### Section 3: Performance metrics

Evaluation of an AI system is most reliably assessed on an unenriched representative sample population. In breast cancer screening, recall rate and cancer detection rate (CDR) are the most informative metrics for evaluating the practical performance of a service. It is worth noting that different screening programmes have different metrics defined and tracked (1,3,4). However, for the purposes of assessing AI in breast cancer screening, the characteristics in Table S3 are always relevant.

**Table S3: An assessment of metrics used for evaluating performance with AI in breast screening**

| Metric | Strengths | Limitations |
|---|---|---|
| Recall Rate | • Common metric tracked in screening programmes<br>• Measurable on the entire sample including unconfirmed cases (not just confirmed positives and confirmed negatives) as not dependent on ground truth definitions<br>• The surrogate for specificity that is relevant for screening practice | • Only meaningful on unenriched unfiltered samples that have the correct screening prevalence |
| Cancer Detection Rate (CDR) | • Common metric tracked in screening programmes<br>• The surrogate for sensitivity that is relevant for screening practice<br>• Intuitive, as it increases for the AI with more IC data available | • Dependent on a positivity ground truth definition<br>• Dependent on the screening programme (screening interval, performance, prevalence) |
| Sensitivity | • A standard diagnostic test metric<br>• Independent of prevalence | • Dependent on a positivity ground truth definition<br>• Not a standardised screening metric<br>• Decreases with more IC data available |
| Specificity | • A standard diagnostic test metric<br>• Independent of prevalence | • Dependent on a negativity ground truth definition<br>• Not a standardised screening metric |

| | | |
|---|---|---|
| Positive Predictive Value (PPV) | • A standard diagnostic test metric sometimes used in screening | • Depending on the PPV definition used (i.e. over all recalls or over sum of true positives and false positives), can mean an upper or lower bound<br>• Dependent on a positivity ground truth definition and may depend on a negativity ground truth definition |
| Arbitration Rate | • Measurable on the entire sample including unconfirmed cases (not just confirmed positives and confirmed negatives) as not dependent on ground truth definitions<br>• Relevant for operational impact in practice | • Implications depend on the nature of the arbitration process which may differ across sites and programmes |

#### Section 4: Determining ground truth, subsample definitions and metrics

Sensitivity, CDR, and PPV were calculated with positives defined as ‘screen-detected positives’ and ‘three-year subsequent cancers’, collectively. Screen-detected positives were screening cases correctly identified by the historical double reader workflow, with a pathology-proven malignancy confirmed by fine needle aspiration cytology (FNAC), core needle biopsy (CNB), vacuum-assisted core biopsy (VACB), and/or histology of the surgical specimen within 180 days of the screening exam.

For the UK sites, ground truth for malignancy was obtained via the NHS National Breast Screening Service (NBSS) database including cancer registry information. In Hungary, confirmation of malignancy was obtained from digital pathology reports in patient health records.

Specificity was calculated on negatives defined as any screening case with evidence of a negative follow-up result that includes a mammography reading at least 1,035 days (i.e. two months less than a three-year screening interval) after the original screening date, with no proof of malignancy in between. PPV, CDR, recall rate, and arbitration rate were calculated on all 275,900 eligible cases. Recall rate, CDR, and arbitration rate were calculated on the whole population, which included confirmed positives, confirmed negatives, and unconfirmed cases (neither confirmed positive nor negative) as this reflects the real-world screening population (see Supplement, Section 3).

Three-year subsequent cancers were defined as a screening case with a pathology-proven cancer arising within 1,095 days following the original screening date and aligned with the definition of interval cancers (IC) for three-year screening interval programmes such as in the UK. The two-year screening interval followed at MK meant that all ICs within the two-year screening interval (‘two-year ICs’), and additional cancers detected at the next screening round, were also included as ‘three-year subsequent cancer’ cases. Recognising the importance of screening interval differences, regional analyses for UK and HU were also performed, using two-year ICs in place of three-year subsequent cancers for HU (see Manuscript, Table 3B).

#### Section 5: Standalone and double reading performance by site and vendor

Table S4 presents performance of standalone AI and the historical first reader by site and vendor. Table S5 presents performance of double reading with and without AI by site and vendor as well.

**Table S4: Performance of standalone AI the historical first reader – by site and vendor.**

| **A) MK / IMS Giotto** | | |
|---|---|---|
| **Performance Metric** | **Historical first reader (%)** | **Standalone AI (%)** |
| On ten-year sample: with incomplete IC data available | | |
| Sensitivity[1] | 70.2 (67.0, 73.2) | 78.3 (75.4, 81.0) |
| Specificity | 95.4 (95.0, 95.7) | 96.1 (95.7, 96.4) |
| On one-year sample: with more complete IC data available | | |
| Sensitivity[1] | 60.0 (51.2, 68.2) | 70.4 (61.9, 77.7) |
| Specificity | 96.5 (95.4, 97.3) | 96.2 (95.0, 97.0) |
| **B) NUH / GE** | | |
| **Performance Metric** | **Historical first reader (%)** | **Standalone AI (%)** |
| On ten-year sample: with incomplete IC data available | | |
| Sensitivity[1] | 77.8 (74.6, 80.7) | 76.9 (73.7, 79.9) |
| Specificity | 97.3 (97.0, 97.5) | 89.6 (89.2, 90.0) |
| On one-year sample: with more complete IC data available | | |
| Sensitivity[1] | 67.2 (58.4, 75.0) | 72.3 (63.6, 79.5) |
| Specificity | 97.2 (96.8, 97.5) | 90.6 (89.9, 91.3) |
| **C) LTHT / Hologic** | | |
| **Performance Metric** | **Historical first reader (%)** | **Standalone AI (%)** |
| On ten-year sample: with incomplete IC data available | | |
| Sensitivity[1] | 81.0 (77.8, 84.0) | 79.9 (76.5, 82.9) |
| Specificity | 95.0 (94.7, 95.3) | 89.2 (88.8, 89.6) |
| On one-year sample: with more complete IC data available | | |
| Sensitivity[1] | 82.8 (73.9, 89.1) | 84.9 (76.3, 90.8) |
| Specificity | 95.7 (95.1, 96.2) | 89.3 (88.5, 90.1) |
| **D) ULH / Siemens** | | |
| **Performance Metric** | **Historical first reader (%)** | **Standalone AI (%)** |
| On ten-year sample: with incomplete IC data available | | |
| Sensitivity[1] | 76.7 (73.3, 79.8) | 77.3 (73.9, 80.4) |
| Specificity | 96.4 (96.1, 96.8) | 89.9 (89.3, 90.5) |
| On one-year sample: with more complete IC data available | | |
| Sensitivity[1] | 70.5 (62.9, 77.1) | 73.1 (65.6, 79.4) |
| Specificity | 97.1 (96.6, 97.6) | 89.7 (88.7, 90.6) |

95% confidence intervals are presented in parentheses.

1. The positive pool for sensitivity includes screen-detected positives and ‘three-year subsequent cancers’ (ie. three-year ICs for the UK plus two-year ICs and additional cancers detected at the next screening round for HU). See Supplement, Section 4 for more details.

---

**Table S5: Performance of double reading with and without AI – by site and vendor.**

| **A) MK / IMS Giotto** | | | |
|---|---|---|---|
| **Performance Metric** | **Historical double reading** | **Double reading (DR) with AI** | **Test outcome for DR with AI[1]** |
| On ten-year sample: with incomplete IC data available | | | |
| Recall rate | 9.2% (9.0, 9.4) | 7.8% (7.7, 8.0) | **Superior** |
| CDR[2] | 7.8 per 1000 (7.2, 8.4) | 7.7 per 1000 (7.2, 8.3) | **Non-inferior** |
| Sensitivity[2] | 78.6% (75.7, 81.3) | 77.7% (74.7, 80.4) | **Non-inferior** |
| Specificity | 94.7% (94.3, 95.0) | 95.8% (95.4, 96.1) | **Superior** |
| PPV[2.3] | 8.5% (7.9, 9.2) | 9.8% (9.2, 10.6) | **Superior** |
| On one-year sample: with more complete IC data available | | | |
| Recall rate | 8.6% (8.0, 9.1) | 7.5% (7.0, 8.0) | **Superior** |
| CDR[2] | 7.7 per 1000 (6.2, 9.6) | 7.7 per 1000 (6.2, 9.5) | **Non-inferior** |
| Sensitivity[2] | 64.8% (56.1, 72.6) | 64.0% (55.3, 71.9) | **Non-inferior** |
| Specificity | 95.8% (94.6, 96.7) | 96.9% (95.8, 97.6) | **Superior** |
| PPV[2.3] | 9.1% (7.4, 11.1) | 10.2% (8.3, 12.5) | **Superior** |
| **B) NUH / GE** | | | |
| **Performance Metric** | **Historical double reading** | **Double reading (DR) with AI** | **Test outcome for DR with AI[1]** |
| On ten-year sample: with incomplete IC data available | | | |
| Recall rate | 2.8% (2.7, 2.9) | 2.8% (2.7, 3.0) | **Non-inferior** |
| CDR[2] | 8.8 per 1000 (8.1, 9.5) | 8.6 per 1000 (7.9, 9.3) | **Non-inferior** |
| Sensitivity[2] | 85.5% (82.7, 87.9) | 83.5% (80.6, 86.1) | **Non-inferior** |
| Specificity | 97.9% (97.7, 98.1) | 97.9% (97.7, 98.1) | **Non-inferior** |
| PPV[2.3] | 31.6% (29.5, 33.7) | 30.4% (28.4, 32.5) | **Non-inferior** |
| On one-year sample: with more complete IC data available | | | |
| Recall rate | 2.8% (2.5, 3.2) | 2.8% (2.5, 3.1) | **Non-inferior** |
| CDR[2] | 8.0 per 1000 (6.5, 9.9) | 7.9 per 1000 (6.4, 9.8) | **Non-inferior** |
| Sensitivity[2] | 73.9% (65.4, 81.0) | 73.1% (64.5, 80.3) | **Non-inferior** |
| Specificity | 98.0% (97.7, 98.3) | 98.1% (97.8, 98.4) | **Non-inferior** |
| PPV[2.3] | 28.3% (23.6, 33.5) | 28.2% (23.5, 33.5) | **Non-inferior** |

| C) LTHT / Hologic | | | |
|---|---|---|---|
| **Performance Metric** | **Historical double reading** | **Double reading (DR) with AI** | **Test outcome for DR with AI[1]** |
| On ten-year sample: with incomplete IC data available | | | |
| Recall rate | 5.1% (4.9, 5.3) | 5.05% (4.9, 5.2) | **Non-inferior** |
| CDR2 | 8.3 per 1000 (7.6, 9.0) | 8.0 per 1000 (7.4, 8.8) | **Non-inferior** |
| Sensitivity[2] | 87.1% (84.2, 89.5) | 84.8% (81.7, 87.4) | **Non-inferior** |
| Specificity | 95.9% (95.7, 96.2) | 96.0% (95.7, 96.3) | **Non-inferior** |
| PPV[2.3] | 16.2% (15.0, 17.5) | 15.9% (14.7, 17.2) | **Non-inferior** |
| On one-year sample: with more complete IC data available | | | |
| Recall rate | 4.3% (4.0, 4.7) | 4.1% (3.8, 4.5) | **Superior** |
| CDR[2] | 7.7 per 1000 (6.2, 9.5) | 7.6 per 1000 (6.1, 9.4) | **Non-inferior** |
| Sensitivity[2] | 88.2% (80.1, 93.3) | 87.1% (78.8, 92.5) | **Non-inferior** |
| Specificity | 96.5% (96.0, 96.9) | 96.6% (96.1, 97.1) | **Non-inferior** |
| PPV[2.3] | 17.7% (14.5, 21.5) | 18.3% (15.0, 22.2) | **Superior** |
| **D) ULH / Siemens** | | | |
| **Performance Metric** | **Historical double reading** | **Double reading (DR) with AI** | **Test outcome for DR with AI[1]** |
| On ten-year sample: with incomplete IC data available | | | |
| Recall rate | 3.6% (3.5, 3.8) | 3.6% (3.4, 3.7) | **Superior** |
| CDR[2] | 9.3 per 1000 (8.6, 10.1) | 9.1 per 1000 (8.4, 9.9) | **Non-inferior** |
| Sensitivity[2] | 85.6% (82.7, 88.1) | 83.4% (80.4, 86.1) | **Non-inferior** |
| Specificity | 97.4% (97.0, 97.7) | 97.5% (97.1, 97.8) | **Non-inferior** |
| PPV[2.3] | 25.7% (23.9, 27.6) | 25.6% (23.7, 27.5) | **Non-inferior** |
| On one-year sample: with more complete IC data available | | | |
| Recall rate | 3.4% (3.1, 3.7) | 3.4% (3.1, 3.7) | **Non-inferior** |
| CDR[2] | 9.0 per 1000 (7.5, 10.7) | 8.8 per 1000 (7.4, 10.5) | **Non-inferior** |
| Sensitivity[2] | 77.6% (70.4, 83.4) | 76.3% (69.0, 82.3) | **Non-inferior** |
| Specificity | 97.6% (97.1, 98.0) | 97.7% (97.2, 98.1) | **Non-inferior** |
| PPV[2,3] | 26.2% (22.4, 30.4) | 26.2% (22.3, 30.4) | **Non-inferior** |

95% confidence intervals are presented in parentheses.

1. All test outcomes were based on the relative difference with a two-sided 95% CI. A 10% margin was used for noninferiority testing (see Statistical Methods for details).
2. The positive pool for CDR, sensitivity, and PPV include screen-detected positives and 'three-year subsequent cancers', which are the standard three-year ICs for the UK (see Supplement, Section 4 for further details).
3. Due to the definition of PPV being over all cases recalled, the figures here represent a lower bound of PPV.

#### Section 6: Further statistical details

Two-sided 95% CIs were used, but for the purposes of testing hypotheses we were only interested in one of the limits (i.e. the limit in whichever direction indicated worse performance), and one side of a two-sided 95% interval is equivalent to calculating with a one-sided 97.5% interval.

For non-inferiority and superiority testing, ratios of proportions were used to calculate relative difference. Since a 10% relative margin was used for non-inferiority testing, this meant that the lower bound of the confidence interval for the ratio of the AI system result to the comparator's result had to be above 0.9 for metrics where a higher result indicates better performance. For superiority testing to pass, the lower bound of the confidence interval of the ratio had to be above 1.

#### Supplementary Information References


1. National Health Institutes England, Public Health England. NHS Breast screening programme screening standards valid for data collected from 1 April 2017. 2021. https://www.gov.uk/government/publications/breast-screening-consolidated-programmestandards/nhs-breast-screening-programme-screening-standards-valid-for-data-collected-from-1april-2017.

2. Burnside ES, Vulkan D, Blanks RG, et al. Association between Screening Mammography Recall Rate and Interval Cancers in the UK Breast Cancer Service Screening Program: A Cohort Study. Radiology. 2018 Jul;288(1):47-54.

3. Radiológiai Szakmai Kollégium (Professional Association of Radiologists, Hungary). Az Egészségügyi Minisztérium szakmai protokollja mammográfiás emlőszűrésről és a korai emlőrák diagnosztikájáról. Egészségügyi Közlöny. 2008.05.28; LVIII/10 (2990-3012)

4. Perry N, Broeders M, de Wolf C, et al. European guidelines for quality assurance in breast cancer screening and diagnosis. Fourth edition—summary document. Annals of Oncology. 2008;19(4):614-22.
